# Validation of a polygenic risk score for Frailty in the Lothian Birth Cohort and English Longitudinal Study of Ageing

**DOI:** 10.1101/2023.04.03.23288064

**Authors:** J P Flint, M Welstead, S R Cox, T C Russ, A Marshall, M Luciano

## Abstract

Frailty is a complex trait. Twin studies and a high-powered Genome Wide Association Study (GWAS) conducted in the UK Biobank have demonstrated a strong genetic basis of frailty. The present study utilized summary statistics from this GWAS to create and test the predictive power of frailty polygenic risk scores (PRS) in two independent samples – the Lothian Birth Cohort 1936 (LBC1936) and the English Longitudinal Study of Ageing (ELSA) aged 67-84 years. Multiple regression models were built to test the predictive power of frailty PRS at five time points. Frailty PRS significantly predicted frailty at all-time points in LBC1936 and ELSA, explaining 2.1% (**β** = 0.15, 95%CI, 0.085-0.21) and 1.6% (**β** = 0.14, 95%CI, 0.10-0.17) of the variance, respectively, at age ∼68/∼70 years (p < 0.001). This work demonstrates that frailty PRS can predict frailty in two independent cohorts, particularly at early ages (∼68/∼70). PRS have the potential to be valuable instruments for identifying those at risk for frailty and could be important for controlling for genetic confounders in epidemiological studies.

---

Frailty is a clinical state commonly associated with ageing and weakening in physiology and risk to stressors ^[1]^. This deterioration often leads to poorer health outcomes in later life, including falls, long-term hospital stays, disability, and mortality ^[2]^. Worldwide, the prevalence of frailty is ∼16% in adults above 60 years old ^[3]^; within the United Kingdom, the prevalence of frailty in over 60-year-olds was estimated to be ∼6.5%, rising substantially for adults over 80 years old and estimated at ∼65% for adults over 90 years old ^[4].^ Given that between 2015 and 2030 the number of people aged 60 across the world is expected to grow from 901 million to 1.4 billion, frailty is now a recognised global health issue – as the population ages, the prevalence of frailty is predicted to rise ^[5]^. Population ageing and the rise in age-related conditions, such as frailty, bring a necessity to use omics and data science to understand the aetiology and mechanisms influencing the development of frailty.

Despite growing evidence that frailty is a public health issue, a universal definition or measurement is yet to be established for frailty ^[6]^. Some studies view frailty as a physical condition that should be considered a medical condition/clinical state - measured, for example, with Fried’s frailty phenotype ^[7]^. Others take a wider definition where frailty is characterised by a reduction in strength, endurance, cognitive and physiological function - measured, for example, with the Frailty Index ^[8]^ - all of which contribute to a decline in independent living and an increased risk of death ^[6]^.

The most recognised predictors of frailty are age and sex ^[9, 10]^. Further predictors include: cognitive, physical, biological, lifestyle and environmental factors, social, sociodemographic and psychological factors ^[11, 12]^. Such factors accumulate across the lifespan and during early life. Despite a multitude of risk factors being identified for frailty, the underlying mechanisms behind such risk factors and the development of frailty have yet to be fully understood, making prediction challenging/imprecise at an individual level ^[13, 14]^. To refine the prediction of traits like frailty the genetic propensity for the manifestation of frailty must be explored. Like many human traits, Frailty Index is partly inherited; twin studies have found that genes explain 30-45% of trait variance ^[1]^.

Developments in molecular genetics allow for more refined prediction of complex traits, such as frailty. For example, Genome-Wide Association Studies (GWAS) explore genetic markers across the genomes of many different people to uncover genetic variations associated with diseases and traits ^[15]^ and explore how these are associated with other biological mechanisms. GWAS reveal that much of the genetic basis for many complex traits come from multiple small effects of thousands of variants ^[15, 16]^. Such genetic associations can allow researchers and clinicians to develop methods to detect, delay and prevent diseases and certain traits. Furthermore, given that frailty has been shown to be reversible ^[17, 18]^, polygenic risk scores (PRS) may not only reveal associations with frailty status, but may also assist in identifying groups of higher risk individuals who could benefit from early intervention ^[19]^.

GWAS have only recently reached large enough sample sizes to uncover reliable and reproducible associations between genetic variants and complex polygenic traits. There have not been many high-powered GWAS studies examining frailty ^[15, 16]^. The largest is a GWAS on 164,510 UK Biobank participants, aged between 60 and 70 years, in which 14 loci were found to be associated with the Frailty Index ^[15]^. 13 of these loci were previously associated with diseases and traits such as depression, smoking, Body Mass Index (BMI), cardiovascular diseases, neuroticism, and Human Leukocyte Antigen Proteins (HLA). The Single Nucleotide Polymorphism (SNP)-based heritability for frailty, measured via the Frailty Index, was 11%, lower than twin-based estimates of 30-45%. However, this is unsurprising given that the SNP based estimate does not include rare genetic variation and structural genetic variation that is captured by twin and family modelling – SNPs index common genetic variation. This high-powered GWAS of frailty points towards genetic determinants linked to cardiovascular health, mental health, and brain functioning ^[15]^. Thus, frailty is a highly polygenic trait, and GWAS are important to understand the underlying biology with potential to robustly predict frailty.

Given the highly polygenic nature of frailty, one method that has become increasingly used to investigate the genetic propensity of diseases and traits is polygenic scoring ^[20]^. This method utilises summary statistics from GWAS, which have examined the associations of millions of SNPs with phenotypes of interest, including in this case, frailty. Weightings (regression coefficients) are then taken for each SNP from GWAS data to create a polygenic risk score for genotyped individuals in an independent sample (participants who are not in the targeted GWAS). This PRS indicates the small cumulative effects contributing to a genetic risk or probability of a higher level of a particular disease or trait ^[20, 21]^. There have been multiple updates in polygenic risk scoring methods to achieve efficient and generalizable results ^[21, 22]^.

PRS can identify individuals at high risk to a certain disease or trait, such as cardiovascular disease ^[20]^. Thus, many researchers have advocated the potential for PRS to be instrumental biomarkers in identifying, predicting, and informing treatment in individuals ^[20, 21, and 22]^. Considering the ageing population, as personalised genomics expands and robust data and methods are applied to understand complex diseases and traits, PRS may be an objective tool in understanding and identifying complex traits like frailty. Furthermore, PRS are used to discriminate environmental from genetic sources of variability, allowing outcomes such as frailty to be better understood.

It is clear that frailty is a multifaceted trait, influenced by many genetic determinants linked to various biological, physical, social, psychological, and environmental traits ^[15, 16, and 23]^. Despite recent findings, there remains a dearth of genetic frailty research. Thus far, no studies have utilised the summary statistics from the most recent frailty GWAS ^[15]^ from the UK Biobank to compute polygenic risk scores in independent samples. This study addresses this gap in the literature by applying the Biobank GWAS summary statistics to two independent cohorts, LBC1936 and ELSA, at five different age bands within the range of 67 to 84. The scores will then be used in multiple linear regression models to predict frailty at five different time points.

## Methods

### Prediction Samples

#### LBC1936

The target data contains the genotypes (genome-wide SNPs) and phenotypic data from 1005 older adults in LBC1936. LBC1936 is an ongoing longitudinal study of older adults living in the community in Edinburgh and surrounding Lothian areas of Scotland, United Kingdom ^[24]^. Individuals were initially recruited based on having been part of the Scottish Mental Survey (1947) and have thus far taken part in 5 waves of testing. The mean age at the first wave was 69.58 (SD = 0.83, n = 1091, 548 males), at the second wave was 72.54 (SD = 0.71, n=866, 448 male), at the third wave was 76.30 (SD = 0.68, n = 697, 360 male), at the fourth wave was 79.38 (SD 0.62, n= 550, 275 male) and at the fifth wave was 82.06 (SD = 0.53, n = 431, 209 male). Ethical permission was approved from the Multi-Centre Research Ethics Committee for Scotland (Wave 1: MREC/01/0/56), the Lothian Research Ethics Committee (Wave 1: LREC/2003/2/29), and the Scotland A Research Ethics Committee (Waves 2, 3, 4 and 5: 07/MRE00/58). Written consent was obtained from participants at each of the waves. The genotypes, collected via blood samples from the majority of participants at wave 1 were processed using stringent quality control measures ^[25]^.

#### ELSA

For ELSA, the target data contained polygenic risk scores and phenotypic data from 5448 adults aged between 67 and 84 (from 9 waves/data collection points in ELSA) living in the community in England ^[26]^. To mirror the format of the LBC1936 waves, the ELSA data was split into five groups based on the same age bands. The mean age in the first group (mirror of LBC1936 Wave 1) was 68.44 (SD = 1.10, n = 3983, 1851 male), in the second group (mirror of LBC1936 Wave 2) was 72.4 (SD = 1.09, n = 3491, 1605 male), in the third group (mirror of LBC1936 Wave 3) was 76.37 (SD = 1.09, n = 2727, 1247 male), in the fourth group (mirror of LBC1936 Wave 4) was 79.95 (SD = 0.76, n = 2020, 874 male), and in the fifth group (mirror of LBC1936 Wave 5) was 82.88 (SD = 0.76, n = 1495, 619 male). Unlike LBC1936, samples in ELSA are refreshed with new participants; therefore, the age groups created for ELSA contained longitudinal participants (with more than one frailty measure across groups) and participants with just one measure in one group. Within the ELSA age groups (1-5), which were created to mirror the format of LBC1936 and validate the PRS prediction, there are individuals who have been tested at different times (from ELSA waves 1-9, which spans from 2002/3 to 2018/19). Supplementary tables S1-S5 show the number of participants, and frailty index means and standard deviations in each age group (1-5) by ELSA wave (1-9), supplementary table S6 shows how many participants have longitudinal measures across the five groups.

Ethics for ELSA have been approved via the South Central – Berkshire Research Ethics Committee (21/SC/0030, 22nd March 2021). Polygenic risk score data was acquired through request to the ELSA genetics team and the phenotypic data was curated at the Advanced Care Research Centre from ELSA data available on the UK data service.

### Predictors

#### Discovery sample

GWAS summary statistics for the largest GWAS to date on frailty were sourced from the European Bioinformatics Institute (EBI) GWAS catalogue ^[27]^. The GWAS included 164,610 individuals (48.5% male) from the UK Biobank aged between 60 and 70 years old. The researchers used the Frailty Index based on 49 self-reported items on a range of physical, social, cognitive, and biological characteristics alongside disabilities and diagnoses.

Quality control was carried out on the GWAS summary statistics. Initial steps included ensuring that the effect and non-effect allele were known to ensure the direction of effect is in the correct direction. Any duplicate and ambiguous SNPs were removed.

### Outcomes

#### Phenotypic data

##### LBC1936

Frailty is measured by the Frailty Index, previously constructed in the LBC dataset ^(28)^. The Frailty Index in the LBC1936 constitutes 30 deficits, including physical, biological, social, psychological, and cognitive deficits, consistent with the Frailty Index in previous research ^[8]^. Deficits were either dichotomised as either 0 (absent) or 1 (present); in some cases, 0.5 was used to represent a partially present deficit or were on a continuous scale (e.g. walking time) on a scale ranging from 0 to 1. For each individual, the number of deficits present was summed and divided by the total number of deficits (30). Scores ranged from 0 to 1 – with higher scores indicating higher frailty.

##### ELSA

The Frailty Index in ELSA was derived from the ELSA dataset following previous work ^[16]^. The index contained 62 deficits and a participant would ascertain a frailty score if data were available for 30 (similar, if not the same deficits as LBC1936) out of the 62 deficits. Despite there being more deficits in the Frailty Index used in ELSA than LBC1936, guidelines indicate that as long as a minimum of 30 deficits are used to cover the relevant domains (disability, disease, cognitive functioning) then differences between number of deficits should not be an issue^[8]^. Due to skewness in the data, the frailty index variable was transformed using a square root transformation. Further, the index was calculated in the same way as the LBC1936 index; both LBC1936 and ELSA followed the same guidelines when creating the index ^[8]^. For LBC1936 and ELSA frailty index scores were standardised to allow comparisons when interpreting the results.

### Covariates

Variation in frailty caused by age and sex (the strongest frailty predictors) were controlled in analysis. Four ancestry principal components for LBC1936 and 10 ancestry principal components for ELSA were also included as covariates to account for population stratification, that is, systematic genetic differences due to ancestry differences – both LBC1936 and ELSA only included participants with European ancestry in genotyping.

### Polygenic risk scores LBC1936

Using the summary statistics from the frailty GWAS ^[15]^ (the base data) and the LBC1936 raw genotype and phenotype data (the target data), PRS were created for 1005 individuals at multiple p-value thresholds (*P*T). Quality control processing was done using the R package QCGWAS and PRS were derived using PRSice (version 2) polygenic software ^[29, 30]^.

### ELSA

PRS for 7223 individuals were provided by the ELSA genetics team and were calculated on genotyped data at multiple *P*T to match the thresholds selected in LBC1936. PRS calculation methods and quality control methods in ELSA can be found in the documentation report ^[31]^. When joined with the phenotypic data 5448 ELSA individuals remained for downstream analysis.

### Statistical analysis

Data preparation details can be found in supplementary materials. Prediction analyses were run using the PRSice 2 software and R studio in R version 2022.7.1.554 ^[32].^

Polygenic risk scores for individuals were calculated with P value thresholds optimised to select the best fitting PRS – the p-value thresholds were between 0.01, 0.05, 0.1, 0.3 and 1. That is, multiple PRS were calculated that included SNPs with association p-values less than the specified threshold and the most predictive of these was retained (93579 SNPs under p-value threshold 1 in LBC1936 and 1349585 SNPs under p-value threshold 1 in ELSA). Clumping was performed to thin SNPs according to the linkage disequilibrium (how correlated SNPs close together are) and p-value. The SNP with the smallest p-value in every 250kb frame was retained, and all SNPs having r^2^ > 0.1 were removed from further analysis.

To explore genetic correlation, multiple linear regression models were built on the PRS against the frailty phenotype and adjusted for age, sex and ancestry principal components. As the PRS were constructed in PRSice for LBC1936, the regression models were also run by the program but built separately in R for the ELSA analysis.

A multiple linear regression was first performed between the covariates (sex, age, and ancestry principal components) and the phenotype (Frailty Index), and constitutes the null model. The PRS is then added as a predictor in the model and re-run (i.e., the full model). The variance explained by the PRS (PRS R^2^) was calculated by subtracting the R^2^ of the null model from that of the full model. ELSA groups were mixed across data collection time points; for example, anybody with a Frailty Index aged 67-70 in group 1 could come from any of the 9 waves of ELSA data collection which can span 16 years. Due to this sampling structure, sensitivity analyses were performed to control for potential cohort effects. This consisted of creating dummy variables for each ELSA wave and adding them as covariates to the regression models.

## Results

The mean scores for frailty and age, at each time point, in the LBC1936 and ELSA are shown in Tables 1 and 2. As expected, mean frailty scores increase with age across the waves. The pairwise correlations of frailty between the waves confirmed that frailty is relatively stable over time (see supplementary tables S7 and S8). Correlations were at a minimum moderate, > 0.5 correlation, and most were strong, Pearson’s *r* > 0.7.

**Table 1.**
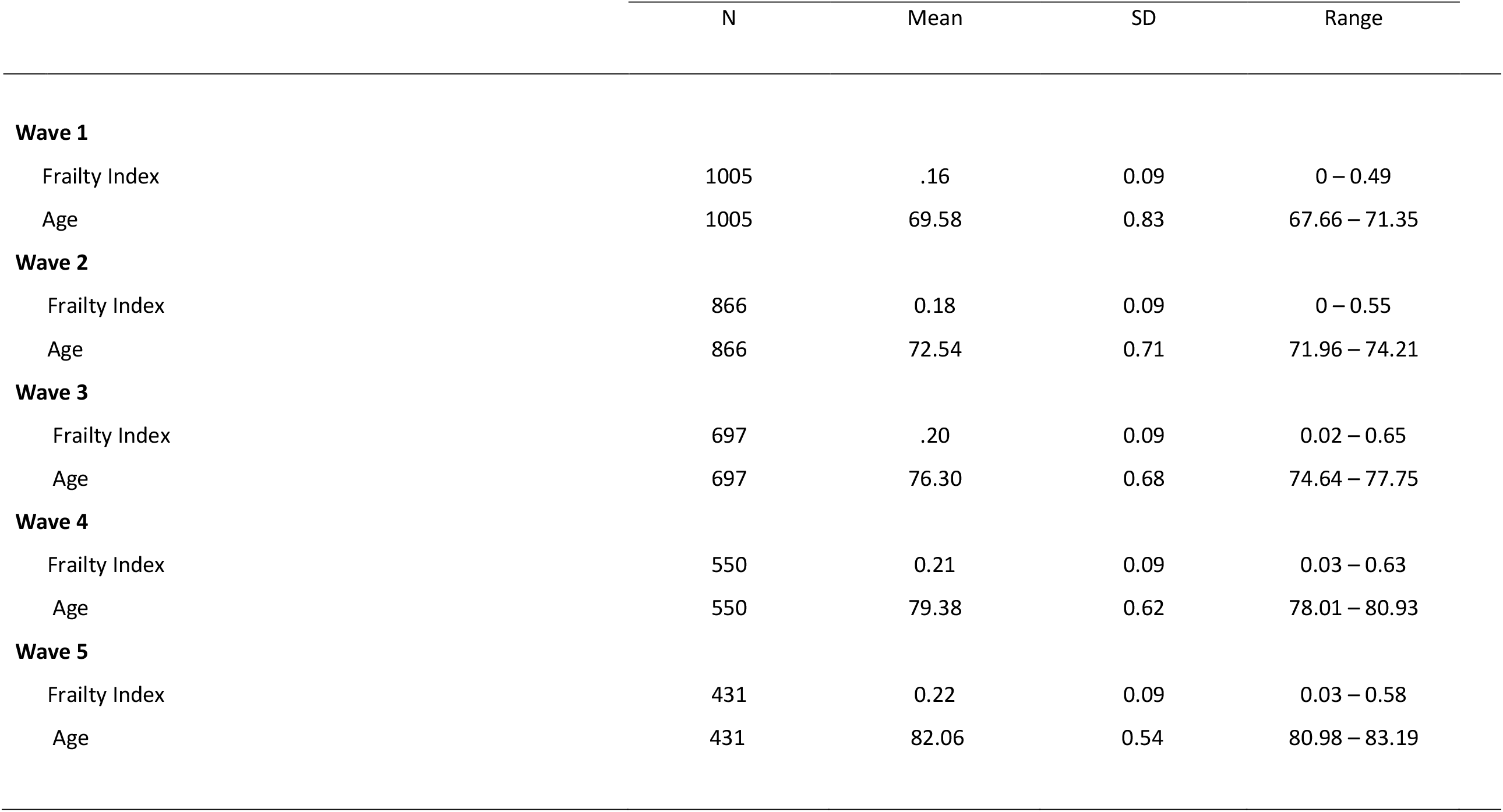
Descriptive statistics for age and raw Frailty Index at each wave in LBC1936

**Table 2.** Descriptive statistics for age and raw Frailty Index at each wave in ELSA

|  | N | Mean | SD | Range |
| --- | --- | --- | --- | --- |
| **Group 1** |  |  |  |  |
| Frailty Index | 3983 | .14 | 0.10 | 0 – 0.68 |
| Age | 3983 | 68.44 | 1.10 | 67.00 – 70.00 |
| **Group 2** |  |  |  |  |
| Frailty Index | 3491 | 0.15 | 0.10 | 0 .007 – 0.66 |
| Age | 3491 | 72.4 | 1.09 | 71.00 – 74.00 |
| **Group 3** |  |  |  |  |
| Frailty Index | 2727 | .18 | 0.11 | 0.007 – 0.69 |
| Age | 2727 | 76.37 | 1.09 | 75.00– 78.00 |
| **Group 4** |  |  |  |  |
| Frailty Index | 2020 | 0.20 | 0.12 | 0.01 – 0.79 |
| Age | 2020 | 79.95 | 0.76 | 79.00 – 81.00 |
| **Group 5** |  |  |  |  |
| Frailty Index | 1495 | 0.22 | 0.12 | 0.01 – 0.74 |
| Age | 1495 | 82.88 | 0.76 | 82.00 – 84.00 |

Figure 1 compares the predictive power of the differing P_T_ in LBC1936, with the bar explaining the most variance representing the best fit. Figure 2 similarly demonstrates the significant relationship at various P_T_ in ELSA. The multiple regression model outputs for each time point can be found in Tables 3 and 4.

**Figure 1.**
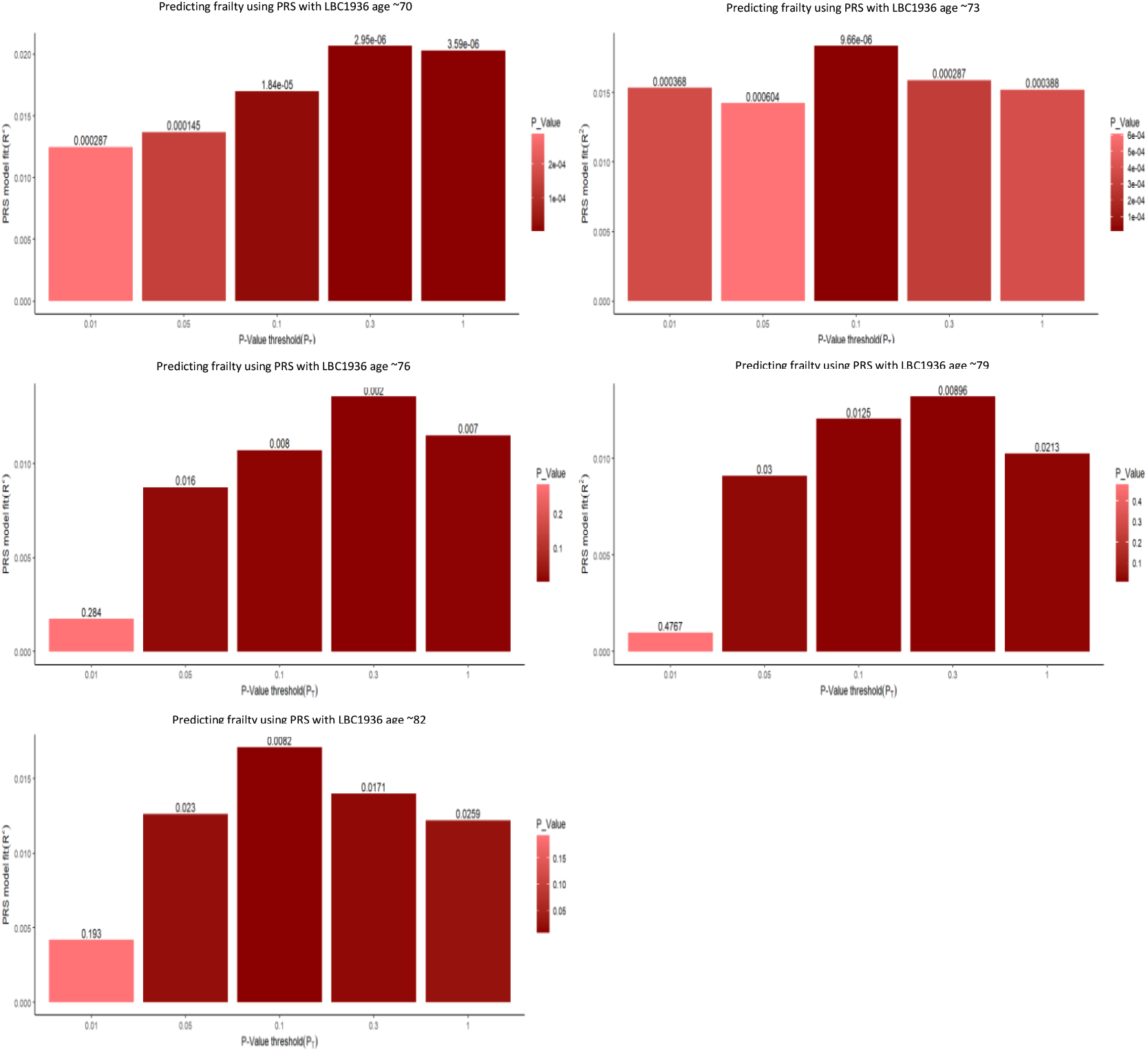
Multiple bar plots, from the multiple regression models, showing the optimal p-value thresholds when predicting frailty using PRS at five time points the LBC1936. The x axis displays the varying different p-value threshold levels. The y axis displays the variance explained by the PGS. The values above the bar are the p-values from the regression output. The darker and taller the bar the stronger the prediction of the frailty PGS.

**Figure 2.**
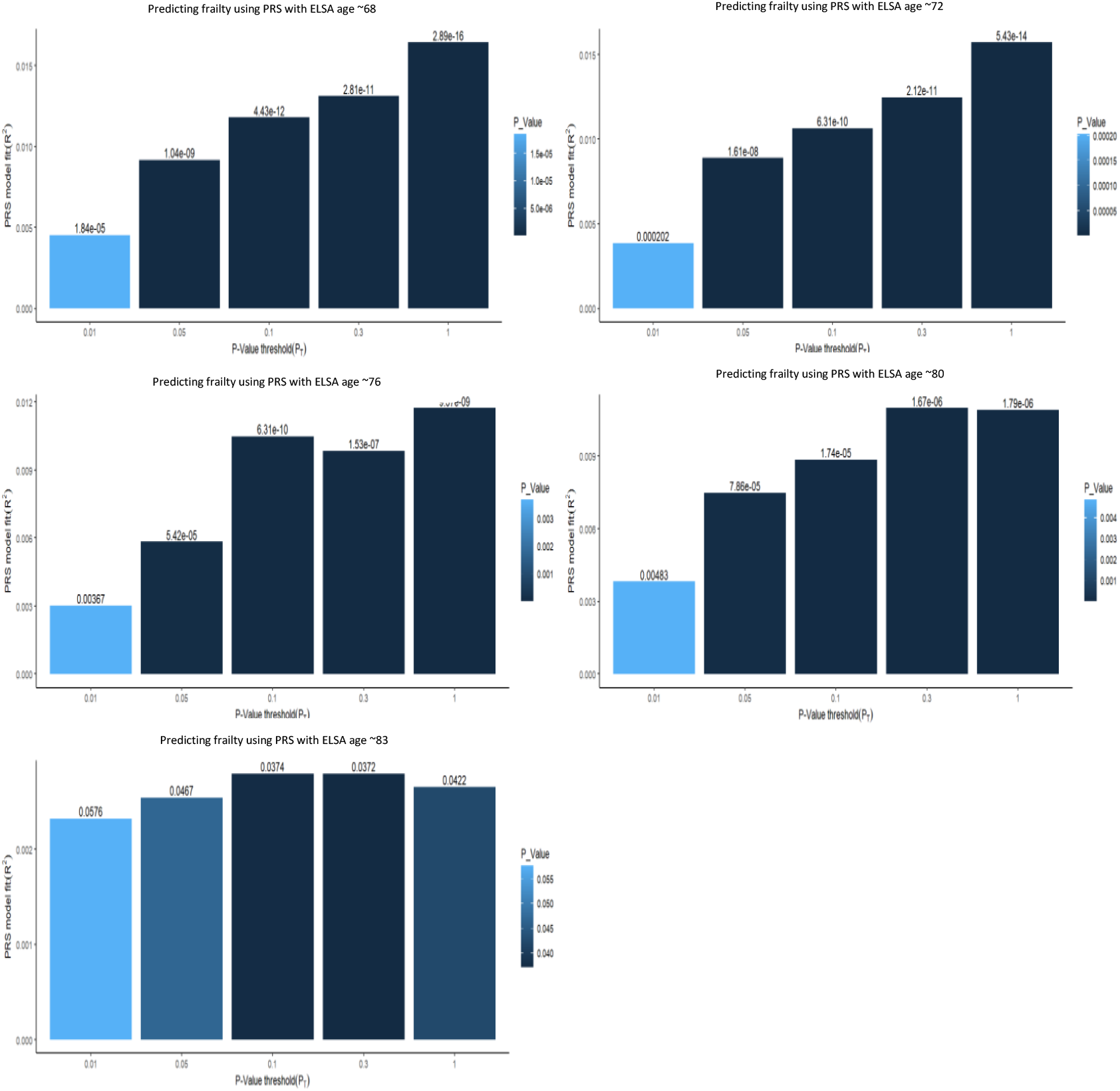
Multiple bar plots, from the multiple regression models, showing the optimal p-value thresholds when predicting frailty using PGS at five time points the ELSA. The x axis displays the varying different p-value threshold levels. The y axis displays the variance explained by the PGS. The values above the bar are the p-values from the regression output. The darker and taller the bar the stronger the prediction of the frailty PRS.

**Table 3.** Results of multiple linear regression analyses showing associations between the optimal frailty PRS and the Frailty Index in the LBC1936.

| Multiple Linear Regression | $\beta$ | SE | $p$ | PRS $R^2$ |
| --- | --- | --- | --- | --- |
| Frailty PRS at ~70 | .15 | 0.03 | <.001 | .021 |
| Frailty PRS at ~73 | .14 | 0.04 | <.001 | .018 |
| Frailty PRS at ~76 | .11 | 0.04 | <.01 | .014 |
| Frailty PRS at ~79 | .11 | 0.04 | <.01 | .013 |
| Frailty PRS at ~82 | .13 | 0.04 | <.01 | .017 |

All analyses controlled for sex and age and population stratification.

**Table 4.** Results of multiple linear regression analyses showing associations between the optimal frailty PRS and the Frailty Index in the English Longitudinal Study of Ageing

| Multiple Linear Regression | $\beta$ | SE | $p$ | PRS $R^2$ |
| --- | --- | --- | --- | --- |
| Frailty PRS at ~68 | .14 | 0.01 | <.001 | .018 |
| Frailty PRS at ~72 | .14 | 0.01 | <.001 | .018 |
| Frailty PRS at ~76 | .12 | 0.01 | <.001 | .013 |
| Frailty PRS at ~80 | .10 | 0.02 | <.001 | .013 |
| Frailty PRS at ~83 | .06 | 0.02 | <.05 | .004 |

All analyses controlled for sex and age, population stratification and ELSA wave.

### LBC1936

At Wave 1, the optimal P_T_ was 0.3, demonstrating the best prediction between frailty PRS and frailty (*p* < .001). As Table 3 shows, the PRS explains 2.1% of variation in frailty within LBC1936 at aged ∼70 and is based on 49325 SNPs. At Wave 2, the optimal P_T_ was 0.1 (*p* < .001) explaining 1.9% of variation in frailty at ∼73 years and is based on 24171 SNPs. At Wave 3, the optimal P_T_ was 0.3 (*p* < .01), explaining 1.4% of variation in frailty at ∼76 years. At Wave 4, the optimal P_T_ was 0.3 (*p* < .05), with the PRS explaining 1.3% of variation in frailty at ∼79 years. At Wave 5, the optimal was 0.1, *p* < 0.1, and the PRS at age ∼82 explains 1.7% of variation in frailty.

### ELSA

For Group 1, the optimal P_T_ was 1, demonstrating the best prediction between frailty PRS and frailty (*p* < .001) explaining 1.6% of variation in frailty at aged ∼68 and based on 1349,585 SNPs. For Group 2, the optimal P_T_ was also 1, with the best prediction between frailty PRS and frailty (*p* < .001) explaining 1.6% of variation in frailty at ∼73 years. For Group 3, the optimal P_T_ was again 1 (*p* < .01), explaining 1.2% of variation in frailty at ∼76 years. For Group 4 the optimal P_T_ was 0.3 (*p* < .05), with the PRS at this wave explaining 1.1% of variation in frailty at ∼79 years and is based on 486,667 SNPs – thus far following similar trends to the variance explained in LBC1936. Finally for Group 5, the optimal P_T_ was 0.1, *p* < 0.1. The PRS explains 0.2% of variation in frailty and is based on 204,805 SNPs.

The findings between LBC1936 and ELSA, apart from at the final time point at Wave/Group 5, were consistent. Figure 3 shows that the standardised coefficients overlap across the waves/groups at each time point across the cohorts, even when the variance drops at the last time point in ELSA – Wave 1 LBC1936 (**β** = 0.15, 95%CI, 0.085-0.21) and Group 1 ELSA (**β** = 0.14, 95%CI, 0.10-0.17); Wave 5 LBC1936 (**β** = 0.13, 95%CI, 0.034-0.23) and Group 5 ELSA (**β** = 0.06, 95%CI, 0.016-0.10).

**Figure 3:**
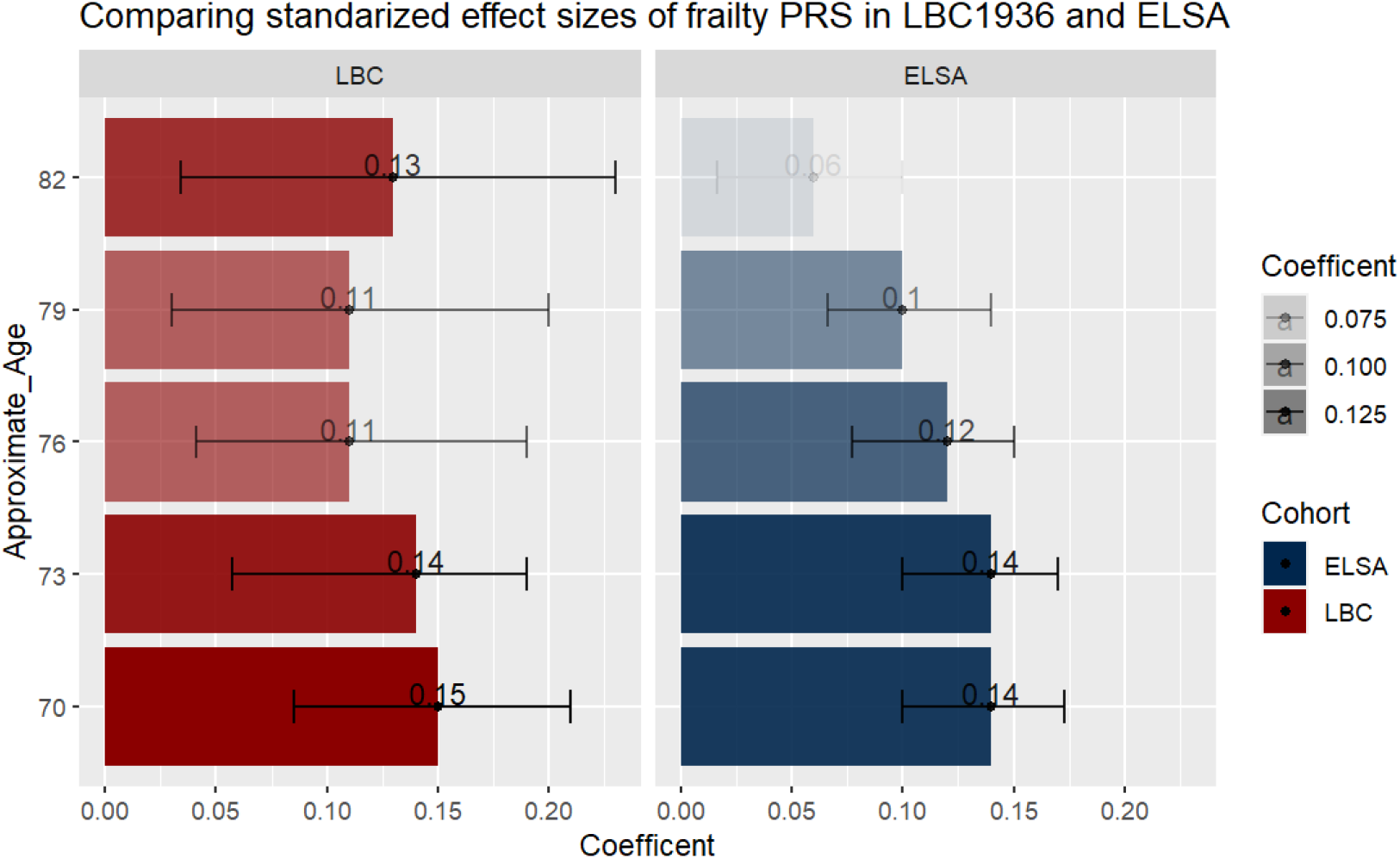
A bar plot comparing the standardized coefficients from the most predictive model at each time point in LBC1936 and ELSA. Error bars represent 95% confidence intervals. The darker the bar the stronger the effect size.

## Discussion

This present study used summary statistics from the most recent and highly powered GWAS on frailty to compute polygenic risk scores (PRS) in an independent sample of community-dwelling older Scottish adults and a nationally representative sample of older adults living in England. Frailty PRS significantly predicted frailty at all five time-points, accounting for the most variance at Wave 1/Age Group 1 (when LBC1936 participants were ∼70 years old and ELSA participants were ∼68 years old); the proportion of variance explained decreased, albeit minimally, across the waves. At the final Wave/Age Group (∼age 82 in LBC 1936 and ∼age 83 in ELSA) the variance explained by frailty PRS had a greater discrepancy between the two cohorts when compared to other ages. Although frailty PRS predicted frailty at all ages, it was most predictive when adults were younger at ∼68/70 years old. This study adds further evidence to the role of genetics in the development of frailty and demonstrates that PRS for frailty can significantly predict frailty outcomes.

Regarding the specific findings, it may have been expected that frailty PRS would explain more variance in adults in the later waves who are approaching and beyond 80 years old given the prevalence of frailty is higher at these ages ^[4].^ But previous research shows that genetic effects on cognitive aging decrease in later life ^[33]^, so it is possible that whereas frailty phenotypic variance increases with age its genetic variance decreases. Our finding that the frailty PRS explained the most variance at the earliest age, when participants were ∼68/70 years, supports such an interpretation. An alternative interpretation of this finding relates to characteristics of the base data (GWAS on frailty) which was performed on adults aged 60 to 70 in the UK Biobank ^[15]^. The age characteristics of the base data were most aligned with the first time point in LBC1936 and ELSA where the mean was ∼68/70 years old. In terms of Frailty Indexing – frailty in the GWAS and at Wave/Age Group 1 in the LBC1936 and ELSA was at a similar mean level. Frailty measured in the GWAS had not yet progressed to higher levels and may not be the most accurate GWAS to use when predicting frailty in older adults, who are at most risk of being frail ^[2, 4]^. Unless there is presence of a chronic condition, frailty would not be screened for at or before age 70 ^[6]^; therefore, measuring frailty between 60-70 years old, as the GWAS did, may limit the utility of the findings. Nonetheless, it still offered significant prediction in our samples across the range of 67 to 84 years.

To understand the findings further, it is useful to understand the frailty score in the GWAS and how that may impact the findings. The Frailty Index mean in the UK Biobank GWAS was 0.12, which most closely resembles the mean frailty levels of .16 and .14 in the respective LBC1936 Wave 1 and ELSA Age Group 1 cohorts. It is possible that frailty measured in a younger group (such as the base data in the UK Biobank) may not necessarily resemble frailty measured at older ages. The example of BMI/weight loss is a good way to illustrate that frailty can mean different things at different ages. In middle age, a relatively lower weight is usually indicative of good health, however in older age a much lower relative weight is indicative of frailty/sarcopenia/poor health ^[34, 35]^ – these age-moderated nuances may not be picked up well in the current GWAS-PRS data. The current study would benefit from research comparing the Frailty Index at an item level with younger and older ages. This would allow researchers to explore the different genetic correlations in frailty at younger ages and at older ages and investigate whether frailty at older ages is tapping the same measure as frailty at younger ages.

Alongside exploring the influence of frailty PRS at increasing ages, the current study has several strengths. The analysis was firstly conducted in LBC1936 and then validated in ELSA. LBC1936 is a unique cohort and is not as representative of the British population when compared to ELSA, thus replicating the analysis with the ELSA cohort increased the validity and generalisability of the findings. Further, the study used p-value thresholding to find the most predictive threshold when creating the polygenic risk scores. Thus, we ensured that the most predictive PRS was used when modelling frailty. The frailty measure in the GWAS and the Frailty Index in both LBC1936 and ELSA used similar indices to create the Frailty Index. If the base and target measures are too dissimilar this can be an issue as it weakens the maximal variance in the target measure that can be explained by the PRS [19, 20].

Despite such strengths, the results should be contextualised within various confines. Firstly, the base data, as previously mentioned, represented frailty at a very early/mild stage ^[36].^ Thus, due to the narrow age examined, the polygenic prediction of frailty at later ages will be biased. Furthermore, participants within the UK Biobank, in which the base data consisted of, are less likely to be obese or inactive, smoke, have lower educational attainment and have fewer health conditions when compared to the general population – the ‘healthy volunteer’ effect ^[37]^. LBC1936 and ELSA are also volunteer studies and are vulnerable to the healthy volunteer effect. The Frailty Index in the base data, LBC1936 and ELSA at ∼70 represented individuals who would be classified as fit or pre-frail – consistent with the idea of healthy participation selection bias ^[38, 39]^. Despite similarities between ELSA and the LBC1936, we imposed group categories in ELSA to represent waves (time points) to mirror the LBC1936 ones. This is a limitation as LBC1936 and ELSA differ in recruitment methods – unlike LBC1936, ELSA recruits and introduces new participants at new time points – this addition of new cohort members (and sampling variation) could explain the lack of variance explained at the last time point in ELSA. Another issue is the reduced sample size from Wave/Age Group 1 to Wave/Age Group 5, and that individuals who remained in the subsequent waves/older ages were healthier on average than those who did not remain in the cohorts ^[38, 39]^. Thus, opportunities were likely missed to study individuals with the highest levels of frailty - this may have been another reason why predictive power decreased at the later waves. In following the GWAS analysis, we deliberately fitted only three covariates (sex, age, and ancestry principal components) in the model given that the frailty PRS is known to genetically correlate with a range of variables which may represent the constituents of frailty; by including them as confounders we would reduce PRS prediction. Future studies could include such covariates alongside their associated PRS to understand if any unique variance in frailty PRS remains following multiple adjustment.

Future studies should continue polygenic prediction of frailty but with more power and refinement to address the challenge of population ageing and support those in and approaching later life. To address the issue of the base data having a narrow age range of 60-70 years, GWAS samples from population representative adults 80+ is needed. This would maximise PRS prediction and allow for more refined identification of those at risk to frailty. This will be challenging as such a GWAS would need to be highly powered with many participants which can prove difficult when attempting to circumvent healthy selection to collect data on older adults, and likely necessitates a meta-analysis approach. A GWAS study conducted on a different measure of frailty - the frailty phenotype – is due to be published ^[40]^. The frailty phenotype takes a physiological approach to measuring frailty, measuring five physical systems weight loss, exhaustion, weakness, slowness when walking and low levels of physical activity ^[7]^. Unlike the Frailty Index it does not feature cognitive elements of frailty and there is a moderate correlation (R = 0.65) between the two ^[6]^. Nonetheless, a future study could test the utility of this PRS in both LBC1936 and ELSA who have measures of the frailty phenotype.

Lastly, the models built in this study contained single polygenic predictors. Despite us showing that single polygenic prediction can be informative when predicting traits like frailty, due to the complexity of frailty, it would add even more value to use a more novel approach such as a multi-polygenic method (MPS) ^[41]^. MPS would increase predictive power via exploiting the combined power of multiple PRS. For example, summary statistics of high-powered GWAS on traits associated with frailty, such as cardiovascular diseases, BMI, and diabetes, could be used to create multiple polygenic risk scores and examine how they perform together in a prediction model to explore the complex nature of frailty.

The present findings have several implications. The summary statistics from the largest GWAS on frailty to date can significantly predict frailty in independent cohorts at multiple time points. This is a starting step in research with complex traits, such as frailty, in utilising genetic data to refine prediction. Polygenic risk scores have the potential to be efficient instruments in detecting genetic liability to a disease or trait and identify these individuals at a point before frailty progresses – a point at which interventions would be effective ^(12,17)^. Furthermore, this study contributes to the small body of research exploring genetic predictors of frailty. These findings support the notion that genetics play an important role in the development of ageing conditions such as frailty and the magnitude of the effect sizes found here are similar to those reported for environmental predictors’ frailty and cognitive ageing ^[42, 43]^. As we tested the associations at five different time points from ages ∼67 to ∼84, this study also supports the importance of an accurate measurement of the Frailty Index – one in which frailty has had the opportunity to progress in both the base and target samples, as this will allow for more refined prediction. Future work could include a high-powered GWAS on advanced frailty, high-powered independent target samples to test frailty PRS in and applying novel methods to utilise multiple PRS in the prediction of frailty.

## Data Availability

The data for the GWAS summary statistics on frailty can be found on: https://www.ebi.ac.uk/gwas/. For the Lothian Birth Cohort 1936 data is available upon request, more information can be found at https://www.lothianbirthcohort.ed.ac.uk/. The English Longitudinal Study of Ageing data was sourced from https://ukdataservice.ac.uk/ and additional PRS were requested via emailing the ELSA team Genetics, ELSA (elsa-project.ac.uk).

## Author Contributions

Concept and Design: J P Flint, M Luciano; Data analysis: J P Flint; Drafting of the manuscript: J P Flint; Critical revision of the manuscript: J P Flint, M Luciano, S R Cox, T C Russ, A Marshall; Administrative technical or material support: M Welstead

## Funding Acknowledgements

This research was funded by the Legal & General Group (research grant to establish the independent Advanced Care Research Centre at the University of Edinburgh). The funder had no role in the conduct of the study, interpretation, or the decision to submit for publication. The views expressed are those of the authors and not necessarily those of Legal and General.

The LBC1936 is supported by the Biotechnology and Biological Sciences Research Council, and the Economic and Social Research Council [BB/W008793/1], Age UK (The Disconnected Mind Project, which also supported MW), the Milton Damerel Trust, and The University of Edinburgh. SRC is supported by a Sir Henry Dale Fellowship jointly funded by the Wellcome Trust and the Royal Society (221890/Z/20/Z). ELSA is funded by the National Institute on Aging (R01AG017644), and by UK Government Departments coordinated by the National Institute for Health and Care Research (NIHR). No editorial service was provided.

## Availability of data and material

The data for the GWAS summary statistics on frailty can be found on: https://www.ebi.ac.uk/gwas/. For the Lothian Birth Cohort 1936 data is available upon request, more information can be found at https://www.lothianbirthcohort.ed.ac.uk/. The English Longitudinal Study of Ageing data was sourced from https://ukdataservice.ac.uk/ and additional PRS were requested via emailing the ELSA team Genetics | ELSA (elsa-project.ac.uk).

