## Supplementary Materials for "Validation of a polygenic risk score for Frailty in the Lothian Birth Cohort and English Longitudinal Study of Ageing"

### Data preparation

In addition to the quality control measures performed on the base and target datasets, the phenotype data (outcome data) had to be cleaned and categorised. Missing data were screened to ensure that any missing values were coded as NA before being inputted into PRSice. To create the polygenic scores in LBC1936, data were then sorted into ten different datasets. Five sets included only the frailty scores of individuals from LBC1936 at each wave – this was then coded into PRSice under the pheno – command. The other five datasets were age, sex and principal components at each wave. Sex and principal components remained consistent, only age differed. The latter five datasets were coded into PRSice under the cov- function.

### PRSice sample script from Wave 1 of LBC1936

The script created to create the polygenic risk scores and run the multiple linear regressions model – this is an example for the PRS and the regressions ran for LBC1936 wave 1.

```
--A1 Effect_allele \
--A2 Other_allele \
--bar-levels 0.05,0.01,0.1,0.5,1 \
--base Basefrailty.txt \
--beta  \
--binary-target F \
--bp Position \
--chr Chromosome \
--pvalue P_value \
```

```
--clump-kb 250kb \
--clump-p 1.000000 \
--clump-r2 0.100000 \
--cov covmerged.txt  \
--cov-col Sex,Age,@C[1-4] \
--cov-factor Sex  \
--fastscore \
--ignore-fid  \
--num-auto 22 \
--out prs.score \
--score avg \
--pheno phenof1.txt \
--seed 270497445 \
--print-snp  \
--snp Markername\
--stat BETA \
--target LBC36.target
```

**Sampling the ELSA data**

ELSA participants, across all five groups, came from 9 collection points (known as waves in ELSA) from Wave 1 2002/2003– to Wave 9 2018/2019. The tables below shows the split of participants across the waves and the mean frailty scores and standard deviations at each wave.

Table S1 showing the spread of group 1 across ELSA wave collection points.

N, Mean, and SD of frailty by ELSA wave age 67-70

| wave | count | mean | sd |
|---|---|---|---|
| 1 | 529 | 0.15 | 0.10 |
| 2 | 395 | 0.13 | 0.09 |
| 3 | 350 | 0.15 | 0.10 |
| 4 | 435 | 0.15 | 0.11 |
| 5 | 396 | 0.14 | 0.10 |
| 6 | 391 | 0.14 | 0.11 |
| 7 | 469 | 0.13 | 0.10 |
| 8 | 467 | 0.14 | 0.11 |
| 9 | 551 | 0.13 | 0.11 |

Table S2 showing the spread of group 2 across ELSA wave collection points.

N, Mean, and SD of frailty by ELSA wave age 71-74

| wave | count | mean | sd |
|---|---|---|---|
| 1 | 436 | 0.17 | 0.10 |
| 2 | 322 | 0.16 | 0.09 |
| 3 | 314 | 0.17 | 0.10 |
| 4 | 401 | 0.15 | 0.09 |
| 5 | 393 | 0.17 | 0.11 |
| 6 | 335 | 0.15 | 0.11 |
| 7 | 343 | 0.15 | 0.11 |
| 8 | 334 | 0.15 | 0.11 |
| 9 | 613 | 0.14 | 0.10 |

Table S3 showing the spread of group 3 across ELSA wave collection points.

N, Mean, and SD of frailty by ELSA wave age 75-78

| wave | count | mean | sd |
|---|---|---|---|
| 1 | 311 | 0.18 | 0.10 |
| 2 | 262 | 0.16 | 0.10 |
| 3 | 265 | 0.18 | 0.11 |
| 4 | 255 | 0.18 | 0.11 |
| 5 | 301 | 0.18 | 0.11 |
| 6 | 321 | 0.16 | 0.11 |
| 7 | 327 | 0.18 | 0.12 |
| 8 | 299 | 0.17 | 0.12 |
| 9 | 386 | 0.17 | 0.11 |

Table S4 showing the spread of group 4 across ELSA wave collection points.

N, Mean, and SD of frailty by ELSA wave age 79-81

| wave | count | mean | sd |
|---|---|---|---|
| 1 | 194 | 0.20 | 0.11 |
| 2 | 198 | 0.19 | 0.10 |
| 3 | 183 | 0.19 | 0.10 |
| 4 | 215 | 0.20 | 0.10 |
| 5 | 209 | 0.21 | 0.12 |
| 6 | 199 | 0.20 | 0.14 |
| 7 | 245 | 0.21 | 0.14 |
| 8 | 291 | 0.20 | 0.12 |
| 9 | 286 | 0.18 | 0.12 |

Table S5 showing the spread of group 5 across ELSA wave collection points.

N, Mean, and SD of frailty by ELSA wave age 82-84

| wave | count | mean | sd |
|---|---|---|---|
| 1 | 126 | 0.21 | 0.11 |
| 2 | 146 | 0.21 | 0.11 |
| 3 | 147 | 0.22 | 0.11 |
| 4 | 153 | 0.22 | 0.12 |
| 5 | 167 | 0.24 | 0.12 |
| 6 | 181 | 0.21 | 0.12 |
| 7 | 155 | 0.22 | 0.14 |
| 8 | 180 | 0.21 | 0.12 |
| 9 | 240 | 0.21 | 0.12 |

Table S6. 1. Numbers of participants who had a previous frailty index measure across the age groups vs participants who only had a single measure at each age group in ELSA.

| Group | Longitudinal participants | Participants with no previous frailty measure | Total |
|---|---|---|---|
| 1/Baseline | 3983 | NA | 3983 |
| 2 | 2812 | 679 | 3491 |
| 3 | 2322 | 405 | 2727 |
| 4 | 1791 | 229 | 2020 |
| 5 | 1345 | 150 | 1495 |

**Correlation matrices**

Table S7. Pairwise correlations of the Frailty Index across the 5 waves in LBC1936

| | **Frailty wave 1** | **Frailty wave 2** | **Frailty wave 3** | **Frailty wave 4** | **Frailty wave 5** |
|---|---|---|---|---|---|
| **Frailty wave 1** | 1*** | | | | |
| **Frailty wave 2** | .80*** | 1*** | | | |
| **Frailty wave 3** | .71*** | .81*** | 1*** | | |
| **Frailty wave 4** | .65*** | .71*** | .79*** | 1*** | |
| **Frailty wave 5** | .57*** | .62*** | .74*** | .79*** | 1*** |

*** all $p < 0.001$

Table S8. Pairwise correlations of the Frailty Index across the 5 waves in ELSA

| | **Frailty wave 1** | **Frailty wave 2** | **Frailty wave 3** | **Frailty wave 4** | **Frailty wave 5** |
|---|---|---|---|---|---|
| **Frailty group 1** | 1*** | | | | |
| **Frailty group 2** | .79*** | 1*** | | | |
| **Frailty group 3** | .71*** | .79*** | 1*** | | |
| **Frailty group 4** | .61*** | .68*** | .75*** | 1*** | |
| **Frailty group 5** | .55*** | .59*** | .64*** | .73*** | 1*** |

*** all $p < 0.001$
